# Burden of SARS-CoV-2 and protection from symptomatic second infection in children

**DOI:** 10.1101/2022.01.03.22268684

**Authors:** John Kubale, Angel Balmaseda, Aaron M Frutos, Nery Sanchez, Miguel Plazaola, Sergio Ojeda, Saira Saborio, Roger Lopez, Carlos Barilla, Gerald Vasquez, Hanny Moreira, Anna Gajewski, Lora Campredon, Hannah Maier, Mahboob Chowdhury, Cristhiam Cerpas, Eva Harris, Guillermina Kuan, Aubree Gordon

**Affiliations:** Department of Epidemiology, University of Michigan School of Public Health, Ann Arbor, MI, USA; Centro Nacional de Diagnóstico y Referencia, Ministry of Health, Managua, Nicaragua; Sustainable Sciences Institute, Managua, Nicaragua; Centro de Salud Sócrates Flores Vivas, Ministry of Health, Managua, Nicaragua; Division of Infectious Diseases and Vaccinology, School of Public Health, University of California, Berkeley, Berkeley, California, USA

**Author notes:** **Corresponding author:** Dr. Aubree Gordon, School of Public Health, University of Michigan, Ann Arbor, MI 48109. Authors contributed equally (John Kubale, Angel Balmaseda).

## Abstract

**Importance:** The impact of the SARS-CoV-2 pandemic on children remains unclear. Better understanding of the burden of COVID-19 among children and their protection against re-infection is crucial as they will be among the last groups vaccinated.

**Objective:** To characterize the burden of COVID-19 and assess how protection from symptomatic re-infection among children may vary by age.

**Design:** A prospective, community-based pediatric cohort study conducted from March 1, 2020 through October 15, 2021.

**Setting:** The Nicaraguan Pediatric Influenza Cohort is a community-based cohort in District 2 of Managua, Nicaragua.

**Participants:** A total of 1964 children aged 0-14 years participated in the cohort. Non-immunocompromised children were enrolled by random selection from a previous pediatric influenza cohort. Additional newborn infants aged ≤4 weeks were randomly selected and enrolled monthly, via home visits.

**Exposures:** Prior COVID-19 infection as confirmed by positive anti SARS-CoV-2 antibodies (receptor binding domain [RBD] and spike protein) or real time RT-PCR confirmed COVID-19 infection ≥60 days prior to current COVID-19.

**Main Outcomes and Measures:** Symptomatic COVID-19 cases confirmed by real time RT-PCR and hospitalization within 28 days of symptom onset of confirmed COVID-19 case.

**Results:** Overall, 49.8% of children tested were seropositive over the course of the study. There were also 207 PCR-confirmed COVID-19 cases, 12 (6.4%) of which were severe enough to require hospitalization. Incidence of COVID-19 was highest among children aged <2 years—16.1 per 100 person-years (95% Confidence Interval [CI]: 12.5, 20.5)—approximately three times that of children in any other age group assessed. Additionally, 41 (19.8%) symptomatic SARS-CoV-2 episodes were re-infections, with younger children slightly more protected against symptomatic reinfection. Among children aged 6-59 months, protection was 61% (Rate Ratio [RR]:0.39, 95% CI:0.2,0.8), while protection among children aged 5-9 and 10-14 years was 64% (RR:0.36,0.2,0.7), and 49% (RR:0.51,0.3-0.9), respectively.

**Conclusions and Relevance:** In this prospective community-based pediatric cohort rates of symptomatic and severe COVID-19 were highest among the youngest participants, with rates stabilizing around age 5. Reinfections represent a large proportion of PCR-positive cases, with children <10 years displaying greater protection from symptomatic reinfection. A vaccine for children <5 years is urgently needed.

**Key Points:** *Question:* What is the burden of COVID-19 among young children and how does protection from re-infection vary with age?

*Findings:* In this study of 1964 children aged 0-14 years children <5 years had the highest rates of symptomatic and severe COVID-19 while also displaying greater protection against re-infection compared to children ≥10 years.

*Meaning:* Given their greater risk of infection and severe disease compared to older children, effective vaccines against COVID-19 are urgently needed for children under 5.

---

The SARS-CoV-2 pandemic has resulted in severe disease and death globally, particularly among the elderly and immunocompromised.^1,2^ Although most pediatric SARS-CoV-2 infections are mild or asymptomatic, severe illness occurs in children, including typical severe respiratory infection and multi-system inflammatory syndrome (MIS-C).^3,4^ Though less common than in adults, studies have suggested that younger children, older adolescents, and children with underlying health conditions more frequently present with symptomatic or severe COVID-19.^5-7^ They may also be more likely to have post-acute sequelae, often referred to as “long covid”, though still less likely compared to adults.^8,9^ However, most of the literature regarding the burden of SARS-CoV-2 in children comes from hospital-based studies, which underestimate the disease burden in children—particularly those with mild infections.^4,6,10-14^ Further, what pediatric community-level evidence does exist is largely from high-income countries and includes few children <5 years.^15-17^As such, despite understanding that differences in infection severity exist between adults and children, the burden and characteristics of SARS-CoV-2 infection among children remain poorly characterized.^3,18^

Understanding the implications of SARS-CoV-2 for children is particularly important, as children will be among the last to be vaccinated. At the time of writing, only one vaccine has been approved in the United States and Europe for children under age 11^19^, and vaccine supplies for many countries— particularly low-and middle-income countries (LMICs)—remain limited.^20^ In fact, as of November 12, 2021, only6.5% of people in low-income countries have received at least one dose.^20,21^ However, even in high-income countries where older children are already being vaccinated, it is critical that we understand the natural history of SARS-CoV-2 in children.

As a consensus has grown around the likelihood of SARS-CoV-2 becoming endemic, researchers have begun exploring what the transition to endemicity might look like so that interventions might be tailored to be more effective. The strength and durability of immune protection against SARS-CoV-2 is perhaps the most important factor in this consideration. A February 2021 study by Lavine et al. suggests that even though sterilizing immunity may wane quickly, if protection against severe infections remains relatively stable, SARS-CoV-2 may become no more severe than the known seasonal human coronaviruses.^22^ However, key uncertainties in the natural history of SARS-CoV-2 infection limit our ability to predict whether this will occur. Of particular importance is the strength and durability of immune protection against illness across the severity spectrum following natural infection in children and whether the increased circulation of variants of concern has impacted the severity of COVID-19 in children. These questions are also of great relevance to future emerging pathogens with pandemic potential. Using data from a community-based prospective pediatric cohort in Managua, Nicaragua, we aimed to assess the burden of infection and disease in the cohort, the strength of protection from symptomatic re-infection, and the relative severity of symptomatic re-infections.

## Methods

### Ethics statement

The study was approved by the Institutional Review Boards of the Nicaraguan Ministry of Health and the University of Michigan. Written informed consent was obtained from a parent/guardian of all participants, and verbal assent was obtained from children aged ≥6 years.

### Study population and sample collection

Participants were from the Nicaraguan Pediatric Influenza Cohort, the methods of which have been described in detail previously.^23^ Briefly, children aged 0-14 years were enrolled when visiting the Health Center Sócrates Flores Vivas (HCSFV), or through home visits, and followed until their 15^th^ birthday or loss to follow-up.

Parents agreed to bring their children to the study clinic (HCSFV) for any illness, and all participants were provided with free medical care for the duration of their participation. Initially, respiratory samples were collected from participants aged 2-14 years presenting with fever/parent-reported fever and at least 1 of the following: cough, sore throat, or rhinorrhea, while respiratory samples were collected from participants aged <2 years who presented with fever/parent-reported fever. However, in June 2020 these criteria were expanded so that samples were also collected from children presenting with loss of taste or smell, rash, conjunctivitis, or fever without a defined focus. Samples were collected using nasal and oropharyngeal flocked plastic swabs. Blood samples were obtained from children upon enrollment and yearly thereafter, in February/March 2020 and February/March 2021. A subset of children had an additional blood sample collected in September/October 2020.

### Laboratory methods

RNA was extracted from respiratory samples (QIAamp Viral RNA Mini Kit, Qiagen) and tested for SARS-CoV-2 via real time RT-PCR.^24^ Blood samples were tested for antibodies against the SARS-CoV-2 receptor-binding domain (RBD) and spike proteins via enzyme-linked immunosorbent assay (ELISA).^25^ Samples were tested for an endpoint using fourfold dilution from 100 until 6400, and the titer for each sample was obtained using the Reed and Muench method.

### Data collection and case definitions

Yearly surveys were administered at the individual and household levels every March/April, collecting data on a wide variety of social and environmental factors. A SARS-CoV-2-specific survey was also completed in September/October 2020 and February/March 2021. Finally, comprehensive, systematic medical consult forms were collected upon each visit to the study health clinic. Data were also collected at follow-up visits that were conducted until the resolution of a child’s illness.

We classified the severity of COVID-19 as: subclinical, mild, moderate, or severe. Participants who reported no symptoms associated with their infection were classified as subclinical. Those with any symptoms excluding difficulty breathing, rapid breathing, or shortness of breath were classified as mild. Moderate COVID-19 was considered illness with associated difficulty breathing, rapid breathing, or shortness of breath. Finally, those requiring hospitalization were classified as having severe COVID-19. A more detailed description of the case definitions used in this analysis can be found in the supplementary materials.

Re-infection with SARS-CoV-2 was defined as a having a positive PCR result after an ELISA-positive result and/or a positive PCR occurring 60+ days after an earlier PCR-positive result. We considered PCR-positive results occurring within 59 days of each other to be from the same infection. For ELISA-positive participants who were not positive by PCR, we utilized surveys conducted in October/November 2020 and February/March 2021 to retrospectively assess COVID-19 illness severity. We observed almost no transmission of influenza or RSV during periods of elevated SARS-CoV-2 transmission, thus participants reporting symptoms that fell within these time periods were assumed to be related to SARS-CoV-2. Participants reporting symptoms between August 1, 2020-February 15, 2021, were considered not SARS-CoV-2-related unless there was a clear epidemiologic link (e.g., confirmed infection in the household at the same time).

### Statistical analysis

We calculated incidence rates for COVID-19 and COVID-19 associated hospitalization using a Poisson distribution to estimate 95% confidence intervals.^26,27^ Protection following SARS-CoV-2 infection was calculated for those with blood samples collected in October 2020 or later. Participants were considered “seropositive” starting the day their first ELISA-positive sample was collected. They were assumed to remain seropositive unless they had a subsequent sample that tested negative by ELISA. We estimated the protection from symptomatic re-infection as 1-Incidence rate ratio. The incidence rate ratio was obtained by fitting a generalized linear model with a Poisson distribution. All analyses were conducted using R version 4.1.1.

## Results

Between March 1, 2020, and October 15, 2021, a total of 1964 children participated in the study, contributing a total of 2733.6 person years, an average of1.4 years per person (Table 1). One hundred and fifty-nine (8.1%) children withdrew from the study or were lost to follow-up. The most common reasons for early withdrawal were failure to appear for their yearly annual sample (44.0%) and inability to find the participant at home (27.7%). Participants recorded 9804 visits to the health center with 1497 (76.2%) children having at least one visit. In all, 1014 (51.6%) participants were infected with SARS-CoV-2 as determined by PCR or ELISA over the course of the study the study.

**Table 1:** Characteristics of study participants Characteristic.

| Characteristic | N=1964 |
| --- | --- |
| Male<sup>a</sup> | 985 (50.2%) |
| Age<sup>b</sup> | 6.9 (4.4) |
| Time in study (person-years)<sup>b</sup> | 1.4 (0.4) |
| Water tap outside house<sup>a</sup> (n = 1716) | 584 (34.0) |
| Number in household<sup>b</sup> (n = 1716) | 7.9 (4.4) |
| <sup>a</sup>N(%) |  |
| <sup>b</sup>Mean(std. dev) |  |

### Incidence of PCR-confirmed COVID-19

Between March 1, 2020, and October 15, 2021, there were a total of 207 PCR-confirmed COVID-19 cases that occurred in 201 (10.2%) participants (Table 2, Figure 1A). While low-level transmission occurred throughout the study, most cases occurred in two distinct waves: March-August 2020 and April-October 2021 (Figure 1A). The overall incidence rate of COVID-19 in the cohort was7.7 cases per 100 person-years (95% Confidence Interval [CI]:6.6,8.8). When examined by age, children <2 years had the greatest incidence of COVID-19, with 16.1 cases per 100 person-years (95% CI: 12.5, 20.5) (Table 2, Figure 1B). Above age 2, the incidence of COVID-19 was substantially lower, but relatively stable, with rates of5.5 (95% CI:3.7,7.9),5.8 (95% CI:4.3,7.6), and6.9 (95% CI:5.3,8.9) cases per 100 person-years among children aged 2-4, 5-9, and 10-14 years, respectively (Table 2, Figure 1B). Female participants had a slightly higher incidence of COVID-19 when compared to males; however, the difference was not significant (Table 2).

**Table 2:**
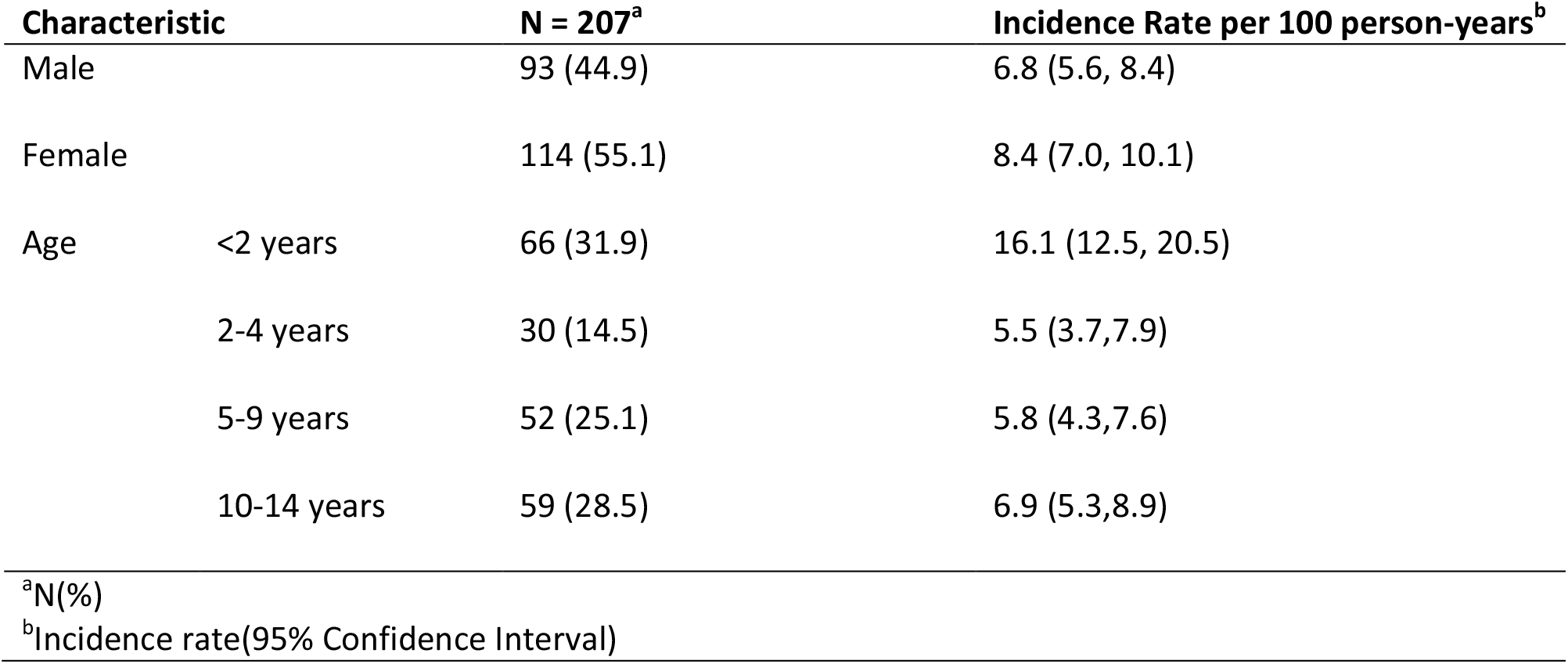
Incidence of Symptomatic SARS-CoV-2.

**Figure 1:**
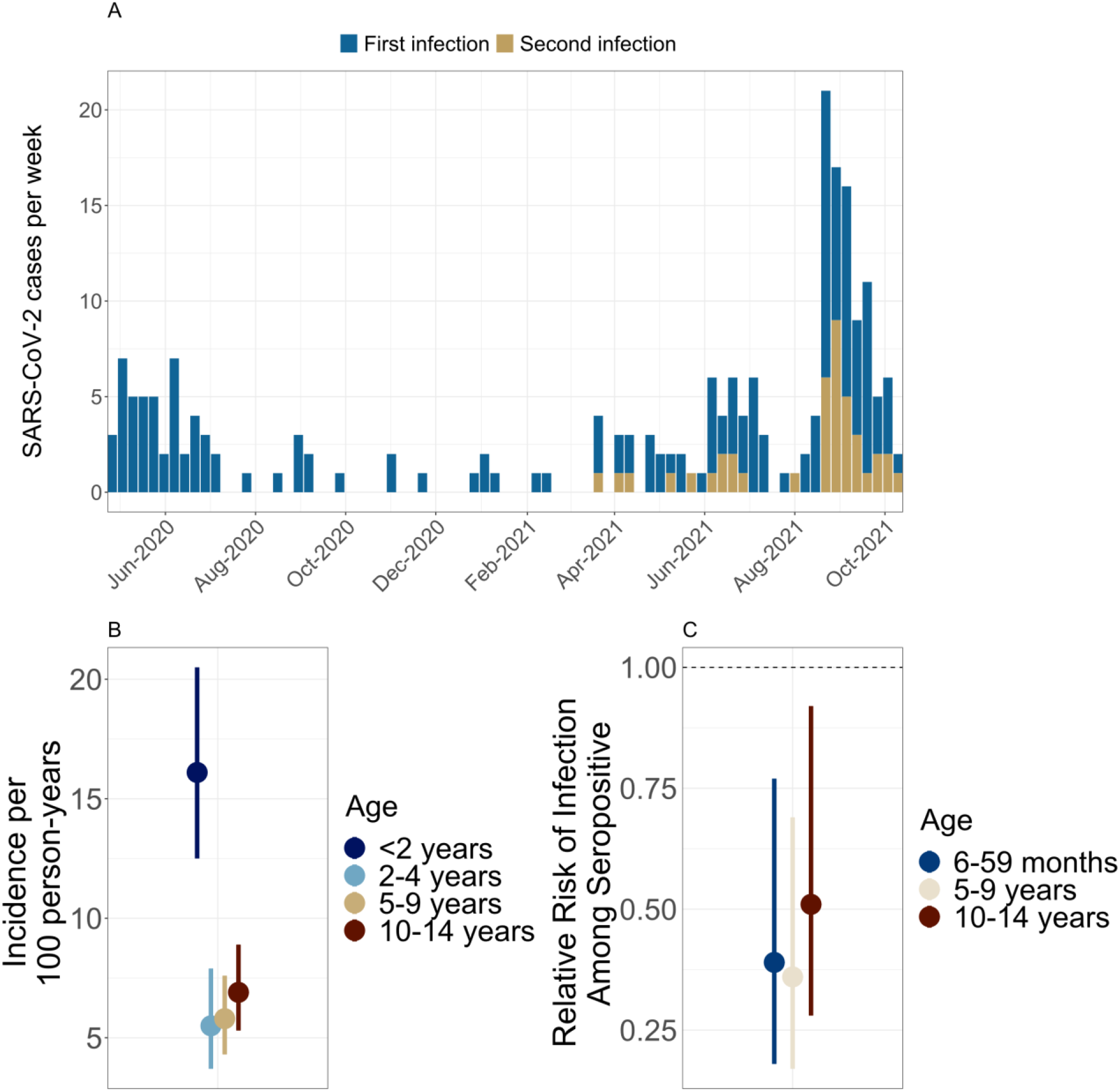
**A) PCR confirmed first and second cases by week over time. B) Incidence rate (per 100 person years) of PCR-confirmed symptomatic SARS-CoV-2 by Age–** 95% confidence intervals obtained using a Poisson distribution. **C) Relative risk of symptomatic infection by seropositivity status** - 95% confidence intervals obtained using a Poisson distribution.

### ELISA-confirmed symptomatic SARS-CoV-2 infection

ELISA results for anti-SARS-CoV-2 antibodies were obtained for 1824 (92.9%) of participants, with 908 (49.8%) being seropositive over the course of the study. Antibody titers were obtained from 877 (96.6%) of seropositive participants. Seroreversion was rare, with 11 (1.2%) previously seropositive participants subsequently testing negative by ELISA. Of these, 10 (90.9%) were >5 years of age, while 1 was <6 months, suggesting that this represented a loss of maternal antibodies. Additionally, among children who seroreverted, only 2 had had illness episodes that were symptomatic (both were mild)—the rest were subclinical. Titers were highest among young children, decreasing between 4-6 years of age before reaching a plateau where they remained relatively stable (Figure 2A). We also saw evidence of relatively rapid waning of maternal antibodies among young infants, with titers steadily decreasing in the first 4 months of life before beginning to increase again (Figure 2B). We observed no meaningful difference in titers by sex (Supplemental Figure 1).

**Figure 2:**
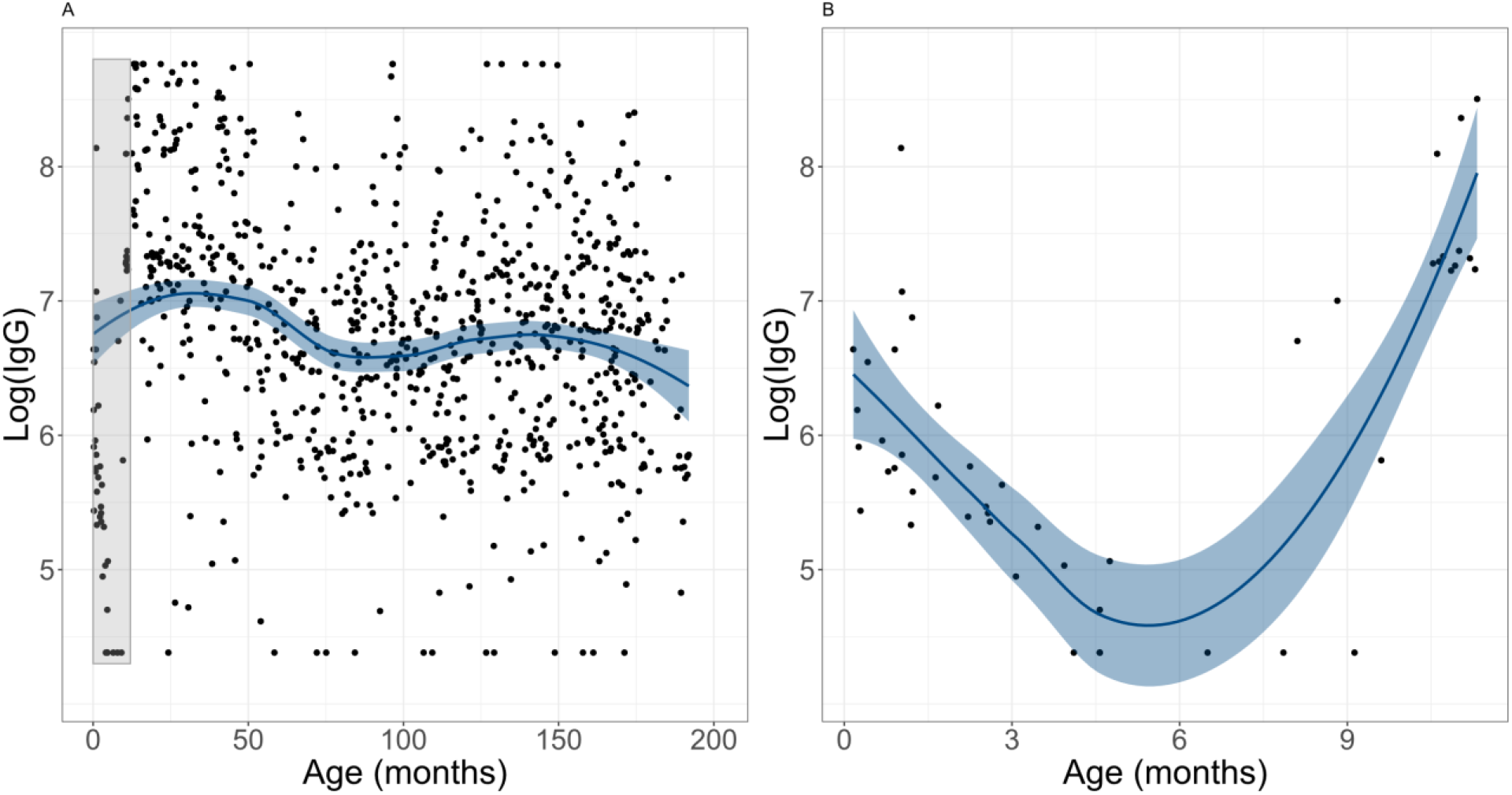
Anti SARS-CoV-2 IgG titers by age -. Panel A shows the log IgG titers of participants by age with a loess smoother and 95% confidence interval to show the overall trend. Panel B shows the exploded view of log titer levels for children aged 0-12 months (the gray shaded region in panel A), with a loess smoother and 95% confidence interval for trend.

### COVID-19 severity

Among PCR-confirmed COVID-19 cases, 12 (5.8%) required hospitalization, resulting in an incidence rate of0.4 (95% CI:0.2,0.8) per 100 person years (Supplemental Table 1). Children aged <2 years had the highest rate of hospitalization associated with COVID-19—2.8 times that of children aged 2-4 years and8.5 times that of children aged 5-9 years (Supplemental Table 1, Supplemental Figure 2A). We did not observe any children aged 10-14 years with COVID-19 who required hospitalization.

### Clinical presentation and long COVID

Children with PCR-confirmed COVID-19 presented with a variety of symptoms, with runny nose (75%), cough (71%), and fever/feverishness (67%) being the most common (Supplemental Table 2, Supplemental Figure 2). While symptoms resolved within four weeks for most children with PCR-confirmed infections, 21 (10.4%) participants had at least one symptom that was present ≥28 days after symptom onset and were classified as having “long COVID”. Of these, 9 (13.6%) were <2 years, 2 (6.7%) were 2-4 years old, 3 (5.8%) were 5-9 years old, and 7 (11.9%) were 10-14 years old. Most participants with long COVID had upper respiratory symptoms that lasted 28 days or longer (14 with runny nose, 7 with cough, 3 with sore throat, and 3 with nasal congestion), while few had long-duration non-specific symptoms (4 still had a fever and 5 still had a headache). One participant under 2 years old still had diarrhea and another under 2 years had rapid breathing.

### Protection from symptomatic second SARS-CoV-2 infection

Symptomatic re-infection with SARS-CoV-2 was relatively common in the cohort, with 41 (19.8%) PCR-confirmed episodes occurring in children with previous SARS-CoV-2 infection detected by PCR or ELISA. All second infections occurred in 2021 when variants of concern, particularly gamma and delta, began actively circulating in Nicaragua.^28^ The relative rate of symptomatic second infection followed a similar pattern to that observed with all COVID-19 cases and associated severe presentations such as hospitalization. Children aged 6 months to 4 years and 5-9 years displayed slightly higher protection from symptomatic second infection at 61% (RR:0.39, 95% CI:0.2,0.8) and 64% (RR:0.36, 95% CI:0.2,0.7) respectively, compared to 49% (RR:0.51, 95% CI:0.3,0.9) among children 10-14 years (Table 3, Figure 1C). Children under 6 months of age at ELISA testing were excluded from the above analysis as they may still have maternal antibody present and thus are not necessarily reinfections. This is consistent with what we observed with antibody titers in the first year of life with a steady decrease in titers in the first six months followed by an increase from 6-12 months (Figure 2B). While moderate and severe secondary cases of COVID-19 did occur, we did not detect any difference in severity between first and second infections (Supplemental Table 3).

**Table 3:** Protection from symptomatic re-infection.

| Characteristic |  | Number of second infections (N=41)<sup>a</sup> | Protection<sup>b</sup> |
| --- | --- | --- | --- |
| Male |  | 14 (34.1) | 62% (RR: 0.38, 95% CI: 0.2,0.7) |
| Female |  | 27 (65.9) | 55% (RR: 0.45, 95% CI: 0.3,0.7) |
| Age | 6-59 months | 10 (24.4) | 61% (RR: 0.39, 95% CI: 0.2,0.8) |
|  | 5-9 years | 11 (26.8) | 64% (RR: 0.36; 95% CI: 0.2,0.7) |
|  | 10-14 years | 20 (48.8) | 49% (RR: 0.51; 95% CI: 0.3,0.9) |
| <sup>a</sup>N(%) |  |  |  |
| <sup>b</sup>Protection calculated as 1-RR |  |  |  |

## Discussion

In this analysis, we assessed the incidence and severity of SARS-CoV-2 infections, as well as protection from symptomatic re-infection, within a prospective, community-based cohort of Nicaraguan children. We observed high rates of infection, with 51.6% of children being infected over the course of the study. While the majority of SARS-CoV-2 episodes were mild or subclinical (Supplemental Table 4), severe illness requiring hospitalization did occur, particularly among children <2 years (Supplemental Table 1). Symptomatic re-infection with SARS-CoV-2 was also common, with 19.8% of PCR-confirmed episodes of SARS-CoV-2 being second infections. This contributed to the lower levels of protection against symptomatic re-infection that we observed compared to previous studies.

Our findings regarding the relative frequency and severity of COVID-19 in children are consistent with those in the literature—specifically that pediatric SARS-CoV-2 infections are common but are largely mild or subclinical.^5-7^ Another study in Nicaragua reported that 37.7% of children aged <15 were seropositive for SARS-CoV-2, substantially lower than the 49.8% we observed in our study.^29^ It was, however, consistent with that estimated from a previously published analysis in a household transmission cohort in the same community where 56.7% of participants were infected.^30^ While over 50% of the cohort were infected in the course of the study, only 207 (19.6%) cases were detected by PCR. This again demonstrates that COVID-19 cases make up only a small proportion of infections in children, and underscores the importance of community-based, prospective studies in accurately characterizing the natural history of pediatric SARS-CoV-2 infection. Previous studies have also indicated that while most children with symptomatic infections observe symptom resolution within a few weeks, some children do experience sustained symptoms broadly described as “long COVID”.^9,15^ Among children with symptomatic SARS-CoV-2 infection, we observed 21 (10.1%) with symptoms that were present ≥28 days after symptom onset, a larger proportion than the4.4% reported by Molteni et al.^15^ However, this difference can likely be attributed to their not including children aged <5 years, as the youngest and oldest age groups in our study displayed the highest frequency of long-COVID. The most common symptoms that persisted beyond 28 days of onset in our study also differed from previous reports, with upper respiratory symptoms being the most common rather than more non-specific symptoms of fatigue and headache reported by others.^9,15^

Given the frequently mild presentation of pediatric SARS-CoV-2 infections, the relative dearth of community-based cohort studies of SARS-CoV-2 infection in children has resulted in a poor understanding of the protection following infection. A previous study in this same community found that protection following infection was similar to that afforded by vaccines^30^; however, this was before variants of concern were widely circulating in Nicaragua and was not powered to assess protection among children under 5. Here, we found that symptomatic second infections were quite common, making up ∼20% of all PCR-confirmed episodes of symptomatic SARS-CoV-2 infection, with children 6-59 months and 5-9 years displaying greater protection against symptomatic second infection (Table 3, Figure 1C). Notably, some of these second infections resulted in illnesses classified as moderate or severe. We were underpowered to sufficiently explore severity of second infections compared to first. Given its implications for pediatric vaccination strategies, the relative severity of second SARS-CoV-2 infections in children should be monitored closely moving forward.^22^

This study has several key strengths. First, as a prospective, community-based pediatric cohort, it provides valuable insight into the burden of symptomatic SARS-CoV-2 infection within a population that is likely to be among the last to be vaccinated—children in low- and middle-income countries. Second, by utilizing well-established systems for sample collection/testing combined with robust data collection, we were able to characterize key aspects of SARS-CoV-2 presentation and severity among children that remain poorly described. Third, by incorporating serologic testing and genetic sequencing, we were able to estimate the protection conferred by natural infection, in the context of variants such as gamma and delta. Fourth, while durability of immune protection against SARS-CoV-2 infection following vaccination is being widely studied, fewer data are available regarding the protection following natural SARS-CoV-2 infection—particularly among children.

This analysis does have several limitations. First, the initial serology samples were collected in February/March 2020—largely before the first wave of SARS-CoV-2 infections in Nicaragua. As such, it is possible that some infections, particularly milder presentations, were missed. It is likely, however, that most of these infections were captured via PCR or by ELISA testing of samples collected in late 2020 and early 2021. Additionally, to assess illness severity of infections detected only by ELISA, we relied on retrospective surveys that may have been subject to recall bias. Fortunately, this is likely to have affected only the mildest of cases as the PCR testing criteria were quite broad—particularly after being expanded in June 2020. Finally, as a community-based, prospective cohort study, we were underpowered to assess the burden of MIS-C and death associated with SARS-CoV-2 infection due to their rarity.

Through this prospective cohort of Nicaraguan children, we were able to assess the impact of SARS-CoV-2 infection on children aged 0-14 years at the community level. We observed high rates of infection, with 51.6% of children having been infected with SARS-CoV-2 over the course of the study. While illness was generally mild, severe illness and prolonged sequelae were observed—most often among children <2 years. Finally, symptomatic re-infection was quite common, with lower protection among children aged >10 years, suggesting important age-associated differences in immune protection following infection.

## Data Availability

Individual-level data may be shared with outside investigators following University of Michigan IRB approval. Please contact Aubree Gordon to arrange for data access.

## Contributions

JTK led the analysis and drafted the manuscript. AB conducted the cohort study and coordinated the molecular and serological testing. GK conducted the cohort study. AG is the Principal Investigator for the Nicaraguan Pediatric Influenza Cohort Study and conceptualized the cohort study and analysis. AMF contributed to the analysis and the drafting/revision of the manuscript. All authors contributed to the manuscript and its revision and approved its submission.

## Data Sharing

Individual-level data may be shared with outside investigators following University of Michigan IRB approval. R code is available on GitHub (https://github.com/jkubale). Please contact Aubree Gordon to arrange for data access.

## Acknowledgements

This work was supported by the National Institute for Allergy and Infectious Diseases (NIAID) at the US National Institutes of Health (grant U01AI088654 and contract HHSN272201400006C) and a grant from Open Philanthropy. We would also like to thank the study participants and their families, the amazing study staff, and the people of Nicaragua. Without them none of this would be possible. We would like to thank Leo Poon for providing the protocol and controls for RT-PCR testing, and Florian Krammer for sharing RBD and Spike constructs as well as technical advice. We are grateful to Janet Smith, Melanie Ohi and their groups at the Center of Structural Biology at the UM Life Sciences Institute for producing proteins and antibodies for the ELISAs.

