## Supplemental Materials for "Burden of SARS-CoV-2 and protection from symptomatic second infection in children"

**Table of Contents:**

Page 3) Case definitions

Page 4) Supplemental Table 1: Incidence of COVID-19 Associated Hospitalization

Page 5) Supplemental Table 2: Symptom presentation of COVID-19 cases by age

Page 6) Supplemental Table 3: Risk of moderate or severe COVID-19 by sero-status

Page 6) Supplemental Table 4: SARS-CoV-2 Illness Severity

Page 7) Supplemental Figure 1: IgG titers by sex

Page 8) Supplemental Figure 2: Incidence rate ratios of COVID-19 associated hospitalization

**Case Definitions:**

| Term | | Definition |
|---|---|---|
| COVID-19 case | | RT-PCR positive for SARS-CoV-2 and no prior positive within previous 59 days |
| Primary episode | | RT-PCR-confirmed case of COVID-19 in participant with no prior positive via PCR or ELISA<br>OR<br>RT-PCR-confirmed case of COVID-19 in participant with prior ELISA positive<br>AND<br>age <6 months at time of ELISA positive |
| Secondary episode | | RT-PCR-confirmed case of COVID-19 in participant with prior ELISA positive<br>AND<br>age ≥ 6 months at time of ELISA positive<br>OR<br>RT-PCR-confirmed case of COVID-19 in participant with prior RT-PCR-confirmed case of COVID-19 occurring >59 days prior. |
| Severity | Subclinical | No reported symptoms |
| | Mild | 1 or more of the following symptoms:<br>• loss of taste<br>• loss of smell<br>• runny nose<br>• cough<br>• headache<br>• sore throat<br>• joint pain<br>• fever<br>• muscle pain<br>• diarrhea<br>• fatigue<br>• rash<br>• stayed in bed<br>• conjunctivitis<br>• congestion<br>• itchy throat<br>• loss of appetite<br>• faintness<br>• tight chest<br>• chest pain |
| | Moderate | Any of the following symptoms:<br>• difficulty breathing<br>• rapid breathing<br>• shortness of breath |
| | Severe | Transfer to hospital |
| Long COVID | | Participants with any symptom persisting for at least 28 days following illness onset for confirmed COVID-19 |

**Supplemental Table 1: Incidence of COVID-19 Associated Hospitalization**

| **Characteristic** | | **N=12** | **Incidence Rate per 100 person-years** |
|---|---|---|---|
| Male | | 7(58.3) | 0.5 (0.2, 1.1) |
| Female | | 5(41.7) | 0.4 (0.1, 0.9) |
| Age | <2 years | 7(58.3) | 1.7 (0.7, 3.5) |
| | 2-4 years | 3(25.0) | 0.6 (0.1, 1.6) |
| | 5-9 years | 2(16.7) | 0.2 (0.03, 0.8) |
| | 10-14 years | 0 | -- |

[a]Confidence intervals calculated using Poisson distribution.

**Supplemental Table 2: Symptom presentation of COVID-19 cases by age**

| Characteristic | <2 years, N = 66[a] | 2-4 years, N = 30[a] | 5-9 years, N = 52[a] | 10-14 years, N = 59[a] | Overall, N = 207[a] |
|---|---|---|---|---|---|
| Loss of taste | 0 (0) | 0 (0) | 3 (5.8) | 8 (14) | 11 (5.3) |
| Loss of smell | 0 (0) | 0 (0) | 3 (5.8) | 11 (19) | 14 (6.8) |
| Runny nose | 48 (73) | 26 (87) | 37 (71) | 44 (75) | 155 (75) |
| Cough | 48 (73) | 23 (77) | 36 (69) | 41 (69) | 148 (71) |
| Headache | 0 (0) | 6 (20) | 24 (46) | 37 (63) | 67 (32) |
| Sore throat | 3 (4.5) | 7 (23) | 26 (50) | 28 (47) | 64 (31) |
| Fever/feverish | 46 (70) | 20 (67) | 35 (67) | 38 (64) | 139 (67) |
| Joint pain | 0 (0) | 1 (3.3) | 3 (5.8) | 14 (24) | 18 (8.7) |
| Muscle pain | 0 (0) | 2 (6.7) | 6 (12) | 15 (25) | 23 (11) |
| Diarrhea | 19 (29) | 3 (10) | 10 (19) | 6 (10) | 38 (18) |
| Vomiting | 5 (7.6) | 0 (0) | 4 (7.7) | 3 (5.1) | 12 (5.8) |
| Fatigue | 0 (0) | 0 (0) | 0 (0) | 1 (1.7) | 1 (0.5) |
| Rash | 1 (1.5) | 1 (3.3) | 0 (0) | 1 (1.7) | 3 (1.4) |
| Stayed in bed | 0 (0) | 0 (0) | 1 (1.9) | 1 (1.7) | 2 (1.0) |
| Conjunctivitis | 0 (0) | 0 (0) | 0 (0) | 0 (0) | 0 (0) |
| Congestion | 30 (45) | 14 (47) | 23 (44) | 23 (39) | 90 (43) |
| Itchy throat | 1 (1.5) | 0 (0) | 3 (5.8) | 5 (8.5) | 9 (4.3) |
| Loss of appetite | 15 (23) | 11 (37) | 17 (33) | 7 (12) | 50 (24) |
| Fainted | 0 (0) | 0 (0) | 0 (0) | 0 (0) | 0 (0) |
| Difficulty breathing | 3 (4.5) | 2 (6.7) | 4 (7.7) | 3 (5.1) | 12 (5.8) |
| Rapid breathing | 4 (6.1) | 1 (3.3) | 2 (3.8) | 0 (0) | 7 (3.4) |
| Shortness of breath | 0 (0) | 0 (0) | 1 (1.9) | 2 (3.4) | 3 (1.4) |
| Chest tightness | 1 (1.5) | 0 (0) | 0 (0) | 1 (1.7) | 2 (1.0) |
| Chest pain | 0 (0) | 0 (0) | 0 (0) | 3 (5.1) | 3 (1.4) |

[a]N(%)

**Supplemental Table 3: Risk of moderate or severe COVID-19 by sero-status**

| | N=10 | Person-years | IRR[a] |
|---|---|---|---|
| Seropositive | 5 | 578.2 | 0.9 (0.3, 3.2) |
| Seronegative | 5 | 515.1 | -- |

[a]Incidence Rate Ratio (95% confidence interval)

**Supplemental Table 4: SARS-CoV-2 Illness Severity**

| Characteristic | | Subclinical | Mild | Moderate | Severe | Total |
|---|---|---|---|---|---|---|
| Female | | 409 (73.6) | 131 (23.6) | 11 (2.0) | 5 (0.9) | 556 |
| Male | | 382 (74.5) | 112 (21.8) | 12 (2.3) | 7 (1.4) | 513 |
| Age | <2 years | 128 (65.3) | 58 (29.6) | 3 (1.5) | 7 (3.6) | 196 |
| | 2-4 years | 148 (79.6) | 32 (17.2) | 3 (1.6) | 3 (1.6) | 186 |
| | 5-9 years | 254 (77.2) | 65 (19.8) | 8 (2.4) | 2 (0.6) | 329 |
| | 10-14 years | 261 (72.9) | 88 (24.6) | 9 (2.5) | 0 (0.0) | 358 |
| Total | | 791 (74.0) | 243 (22.7) | 23 (2.2) | 12 (1.1) | 1069 |

[1] Values correspond to N(%)

**Supplemental Figure 1: Anti SARS-CoV-2 IgG titers by sex**

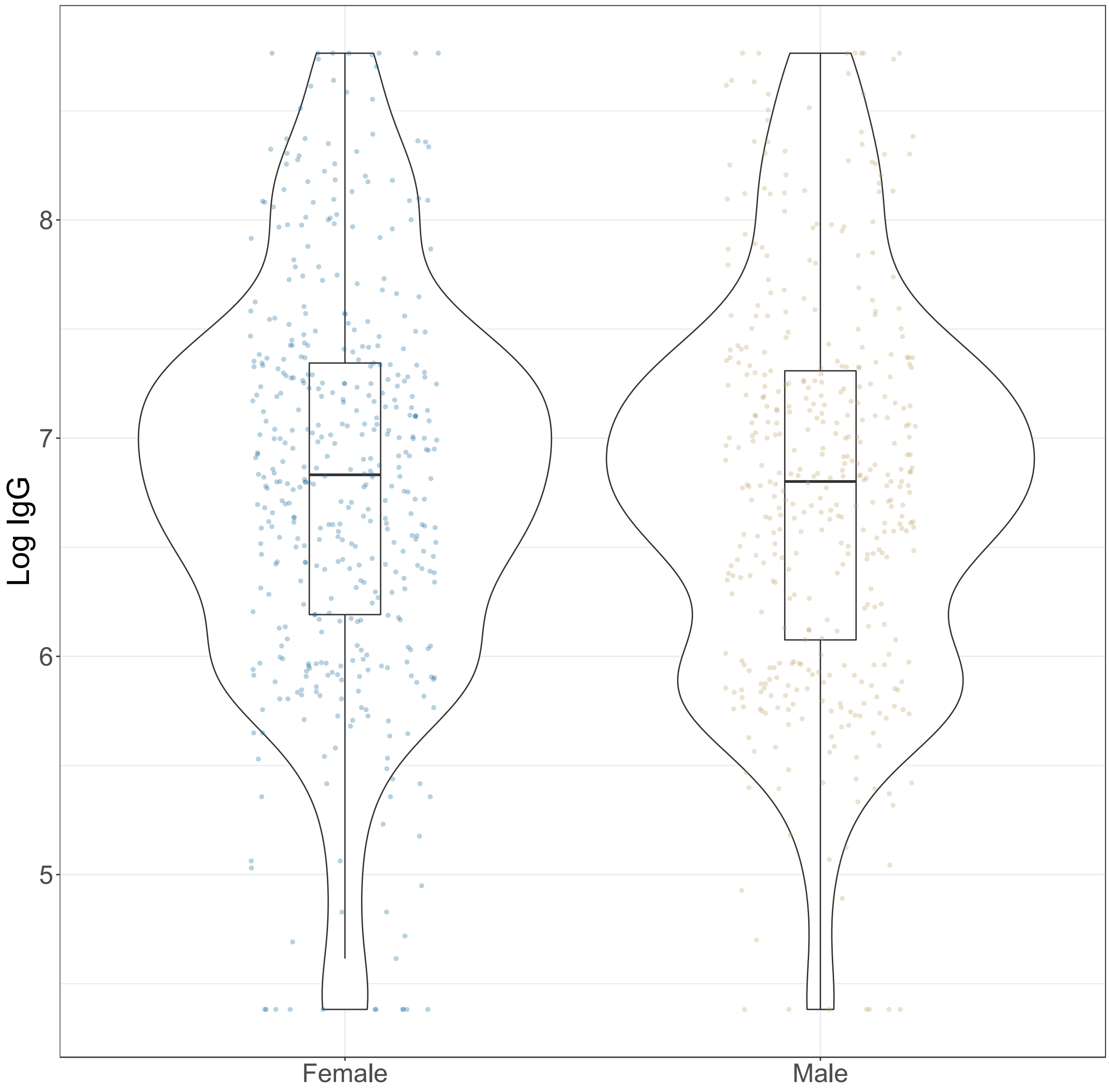

**Supplemental Figure 2: Incidence rate ratios of COVID-19 associated hospitalization**

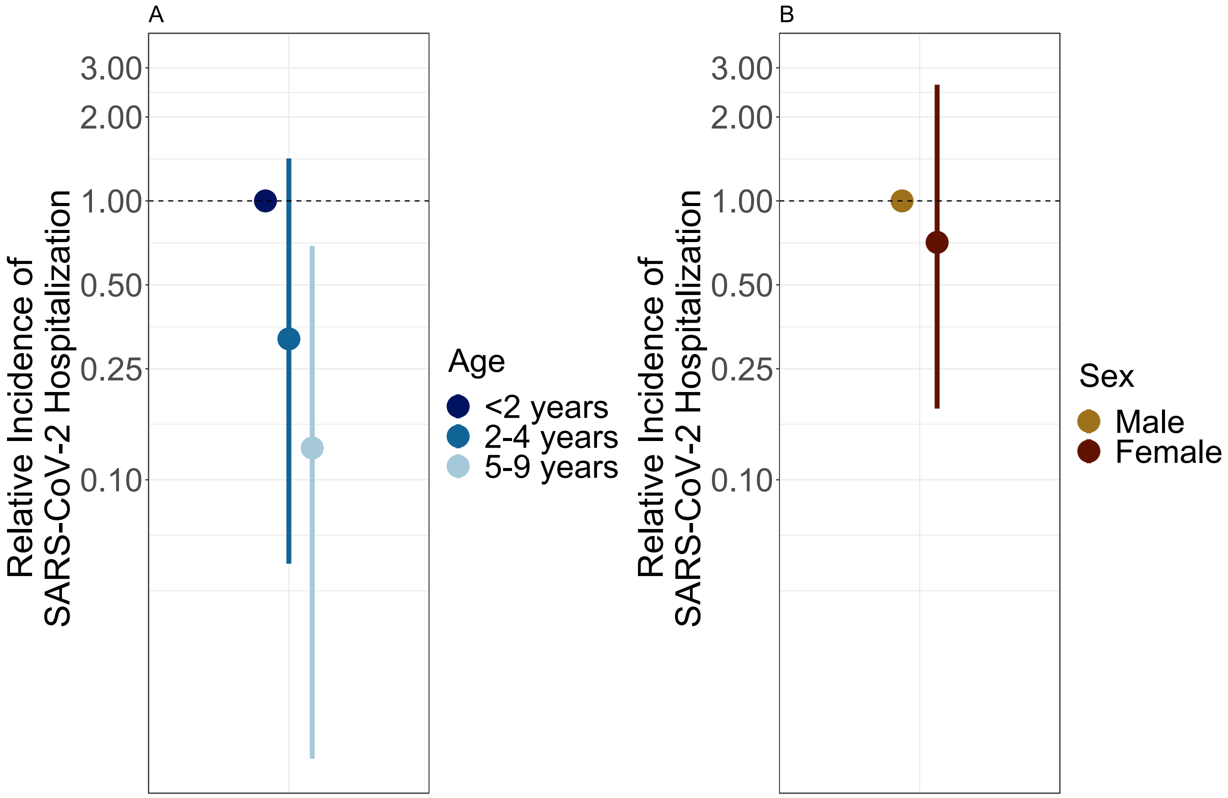
